# International Multicenter Study Comparing Cancer to Non-Cancer Patients with COVID-19: Impact of Risk Factors and Treatment Modalities on Survivorship

**DOI:** 10.1101/2022.08.25.22279181

**Authors:** Issam Raad, Ray Hachem, Nigo Masayuki, Tarcila Datoguia, Hiba Dagher, Ying Jiang, Vivek Subbiah, Bilal Siddiqui, Arnaud Bayle, Robert Somer, Ana Fernández Cruz, Edward Gorak, Arvinder Bhinder, Nobuyoshi Mori, Nelson Hamerschlak, Samuel Shelanski, Tomislav Dragivich, Yee Elise Vong Kiat, Suha Fakhreddine, Pierre Abi Hanna, Roy F. Chemaly, Victor Mulanovich, Javier Adachi, Jovan Borjan, Fareed Khawaja, Bruno Granwehr, Teny John, Eduardo Yepez Guevara, Harrys Torres, Natraj Reddy Ammakkanavar, Marcel Yibirin, Cielito C Reyes-Gibby, Mala Pande, Noman Ali, Raniv Dawey Rojo, Shahnoor M Ali, Rita E Deeba, Patrick Chaftari, Takahiro Matsuo, Kazuhiro Ishikawa, Ryo Hasegawa, Ramón Aguado-Noya, Álvaro García-García, Cristina Traseira Puchol, Dong-Gun Lee, Monica Slavin, Benjamin Teh, Cesar A Arias, Data-Driven Determinants for COVID-19 Oncology Discovery Effort (D3CODE) Team, Dimitrios P. Kontoyiannis, Alexandre E. Malek, Anne-Marie Chaftari

## Abstract

**Background:** In this international multicenter study we aimed to determine the independent risk factors associated with increased 30-day mortality and the impact of novel treatment modalities in a large group of cancer and non-cancer patients with COVID-19 from multiple countries.

**Methods:** We retrospectively collected de-identified data on a cohort of cancer and non-cancer patients diagnosed with COVID-19 between January and November 2020, from 16 international centers.

**Results:** We analyzed 3966 COVID-19 confirmed patients, 1115 cancer and 2851 non-cancer patients. Cancer patients were more likely to be pancytopenic, and have a smoking history, pulmonary disorders, hypertension, diabetes mellitus, and corticosteroid use in the preceding two weeks (p≤0.01). In addition, they were more likely to present with higher inflammatory biomarkers (D-dimer, ferritin and procalcitonin), but were less likely to present with clinical symptoms (p≤0.01). By multivariable logistic regression analysis, cancer was an independent risk factor for 30-day mortality (OR 1.46; 95% CI 1.03 to 2.07; p=0.035). Older age (≥65 years) was the strongest predictor of 30-day mortality in all patients (OR 4.55; 95% CI 3.34 to6.20; p< 0.0001). Remdesivir was the only therapeutic agent independently associated with decreased 30-day mortality (OR 0.58; CI 0.39-0.88; p=0.009). Among patients on low-flow oxygen at admission, patients who received remdesivir had a lower 30-day mortality rate than those who did not (5.9% vs 17.6%; p=0.03).

**Conclusions:** Cancer is an independent risk factor for increased 30-day all-cause mortality from COVID-19. Remdesivir, particularly in patients receiving low-flow oxygen, can reduce 30-day all-cause mortality.

**Condensed Abstract:** In this large multicenter worldwide study of 4015 patients with COVID-19 that included 1115 patients with cancer, we found that cancer is an independent risk factor for increased 30-day all-cause mortality. Remdesivir is a promising treatment modality to reduce 30-day all-cause mortality.

## INTRODUCTION

The COVID-19 pandemic has challenged the health care system worldwide and has spread to more than 200 countries, causing hundreds of millions confirmed cases and several million deaths.^1^

Data from multiple studies have shown consistently that older age and comorbidities such as cardiovascular disease, diabetes mellitus (DM), hypertension, and chronic obstructive pulmonary disease (COPD)have been associated with severe illness and increased mortality.^2^ Several studies on COVID-19 mortality suggested that cancer patients had poor outcomes.^3-^ Many of these studies included only cancer patients and did not have a comparator group of non-cancer patients.^3-7^ Other studies compared COVID-19 mortality between cancer and non-cancer patients and found that cancer patients had worse outcomes.^9-13^ However, all of these comparative studies included a relatively small number of cancer patients and were restricted to a particular country, which limits their generalizability to cancer patients worldwide.

Given that many of the therapeutic studies on COVID-19 were conducted in non-cancer patients and given the poor outcomes of COVID-19 reported in cancer patients,^9-18^ we aimed to conduct a large multicenter study to compare the impact of these various treatments on the outcome in cancer versus non-cancer patients.

Therefore, given the global spread of the COVID-19 pandemic and the worldwide prevalence of cancer, we undertook this international initiative that included 16 centers from five continents to study and compare the clinical course, risk factors and treatment modalities impact on outcomes in cancer versus non-cancer patients with COVID-19 on a worldwide basis.

## Methods

### Study design and participants

This was a retrospective international multicenter study that included all patients diagnosed with COVID-19 by real-time polymerase chain reaction (RT-PCR) for severe acute respiratory syndrome coronavirus 2 (SARS-CoV-2) at the site or an outside facility between January 4, 2020, and November 15, 2020.

The study involved 16 centers from 9 countries, eight centers in the United States and one each in Australia, Brazil, France, Japan, Lebanon, Singapore, South Korea, and Spain. Patients were divided into two groups: non-cancer and cancer patients diagnosed or treated within a year before the diagnosis of COVID-19.

### Multicenter collaboration and data collection

The University of Texas MD Anderson Cancer Center was the coordinating center that designed the study, built the electronic case report form and collected de-identified patient information from all participating centers using the secure Research Electronic Data Capture platform. We reviewed each patient’s electronic hospital record and collected all needed data. This study was approved by the institutional review board at MD Anderson Cancer Center and the institutional review boards of the collaborating centers. A patient waiver of informed consent was obtained. The follow-up period was defined as 30 days after a diagnosis of COVID-19.

Neutropenia was defined as an absolute neutrophil count <500 cells/mL. Lymphopenia was defined as an absolute lymphocyte count <500 cells/mL.

### Treatment modalities

For each patient, data were collected on COVID-19 treatments, including antimicrobial therapy and potential antiviral therapy. We evaluated patient’s oxygen requirement: low-flow was defined as oxygen supplementation of ≤ 6 liters/minute through a nasal cannula or facemask, and high-flow included all other modalities of oxygen supplementation, including mechanical ventilation.

### Outcomes measures

The primary outcome of interest was 30-day mortality. The secondary outcomes included mechanical ventilation, progression to lower respiratory tract infection, co-infection, and hospital readmissions within 30 days after COVID-19 diagnosis.

### Statistical analysis

Patient characteristics and outcomes were compared between COVID-19 patients with and without cancer. Categorical variables were compared using chi-square or Fisher’s exact test, and continuous variables compared using Wilcoxon rank sum test. Kaplan-Meier curves were estimated and compared using a log-rank test. A multivariable logistic regression model was used to identify the independent predictors of 30-day mortality in all patients and in cancer patients and non-cancer patients among the following factors: patients’ demographic and clinical characteristics, medical history, laboratory findings at diagnosis, and treatment. In cancer patients, the mortality rates of patients with different types of cancer were estimated and compared. Lastly, the impacts on mortality of certain therapeutic agents and their timing of administration were evaluated using chi-square or Fisher’s exact test. All statistical tests were two-sided with a significance level of 0.05. The analyses were performed using SAS version 9.3 (SAS Institute Inc., Cary, NC).

## RESULTS

A total of 4015 patients diagnosed with COVID-19 by PCR were included in the study. After excluding 18 patients who had missing demographics and 31 patients younger than 18 years, we evaluated 3966 COVID-19 patients: 2851 non-cancer and 1115 cancer patients.

### Demographics and clinical characteristics of patients with and without cancer

Patient characteristics are presented in Table 1. Compared to non-cancer patients, cancer patients were older (median age, 61 vs 50 years; p<0.0001); more likely to have a smoking history (38% vs 17%; p<0.0001), pulmonary disorder (27% vs 21%; p<0.001), hypertension (49% vs 36%, p<0.0001), DM (27% vs 23%, p=0.01), or corticosteroid use in the two weeks preceding COVID-19 diagnosis (17% vs 4%, p<0.0001); and less likely to present with clinical symptoms, including cough (46% vs 65%; p<0.0001), fever (45% vs 66%; p<0.0001), and shortness of breath (35% vs 48%; p<0.0001).

**Table 1.**
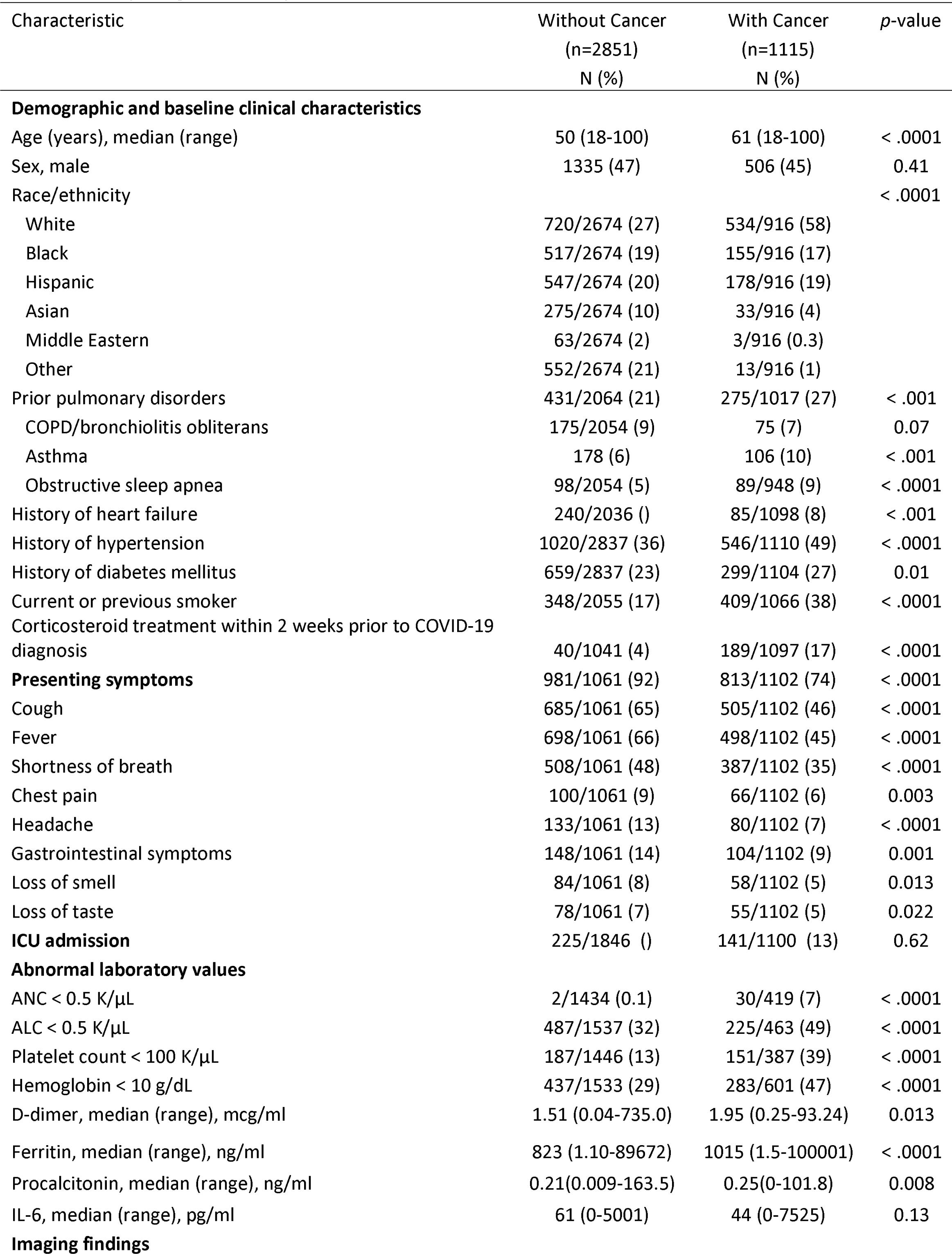
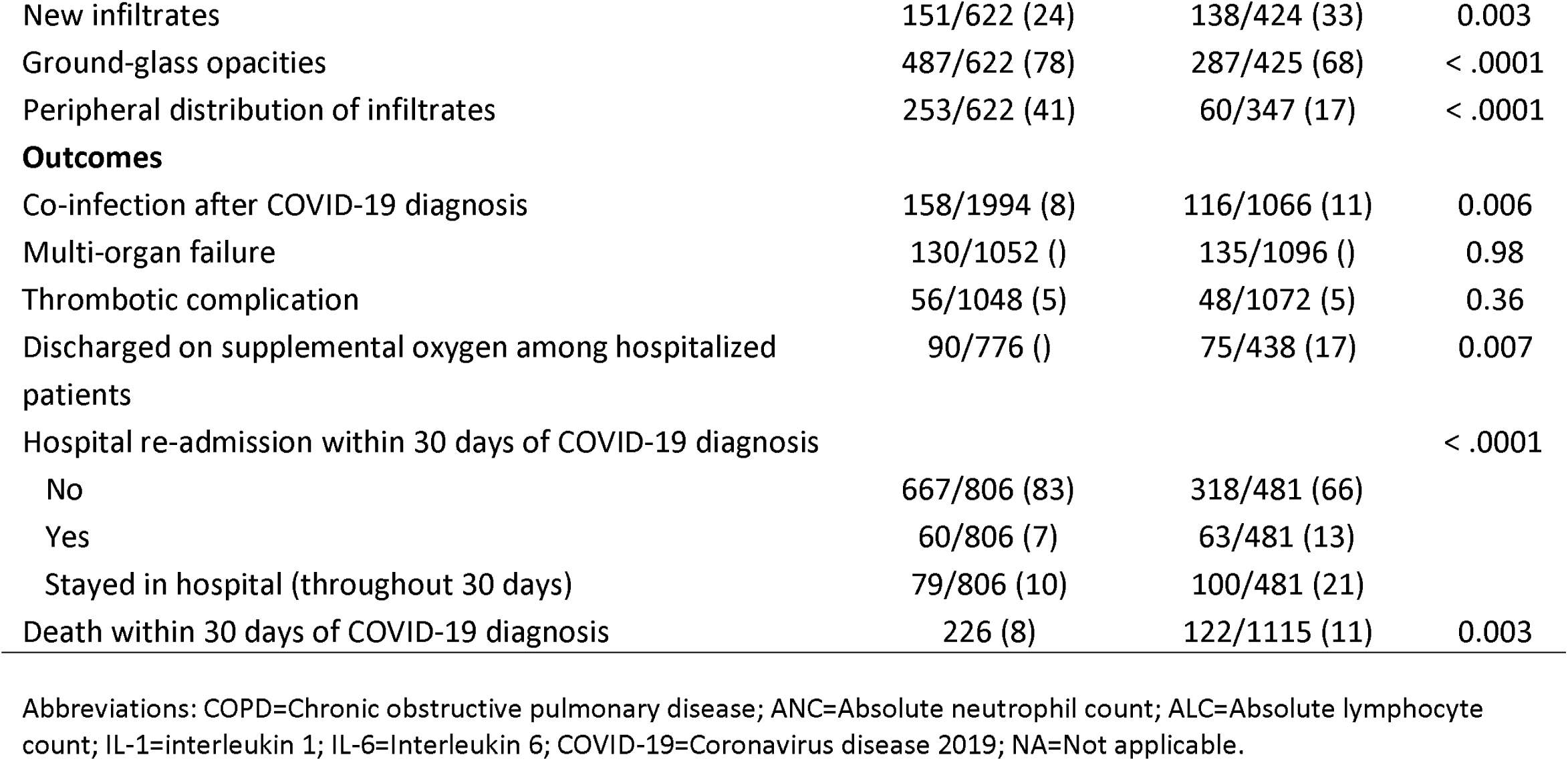
Comparing COVID-19 patients with and without cancer

Compared to non-cancer patients, cancer patients were more likely to present with neutropenia (7% vs 0.1%; p<0.0001), lymphocytopenia (49% vs 32%; p<0.0001), thrombocytopenia (39% vs 13%; p<0.0001), and anemia (47% vs 29%; p<0.0001) and had higher median levels of inflammatory biomarkers, including D-dimer, ferritin, and procalcitonin. On imaging studies (computed tomography), cancer patients were less likely than non-cancer patients to present with ground-glass opacities (68% vs 78%; p<0.0001) or peripheral distribution of the infiltrates (17% vs 41%; p<0.0001).

### Treatment and outcomes of patients with and without cancer

The rate of COVID-related hospital admission was higher in the non-cancer patients (64% vs 57%; p<0.001). Co-infections occurred more frequently in cancer patients (11% vs 8%; p=0.006). Among hospitalized patients, cancer patients were more likely than non-cancer patients to be discharged on supplemental oxygen (17% vs %; p=0.007). Likewise, the rate of hospital readmission within 30 days was higher in cancer patients (13% vs 7%; p<0.0001). Furthermore, the mortality rate within 30 days was significantly higher in cancer patients (11% vs 8%; p=0.003) (Table 1 and Figure 1).

**Figure 1.**
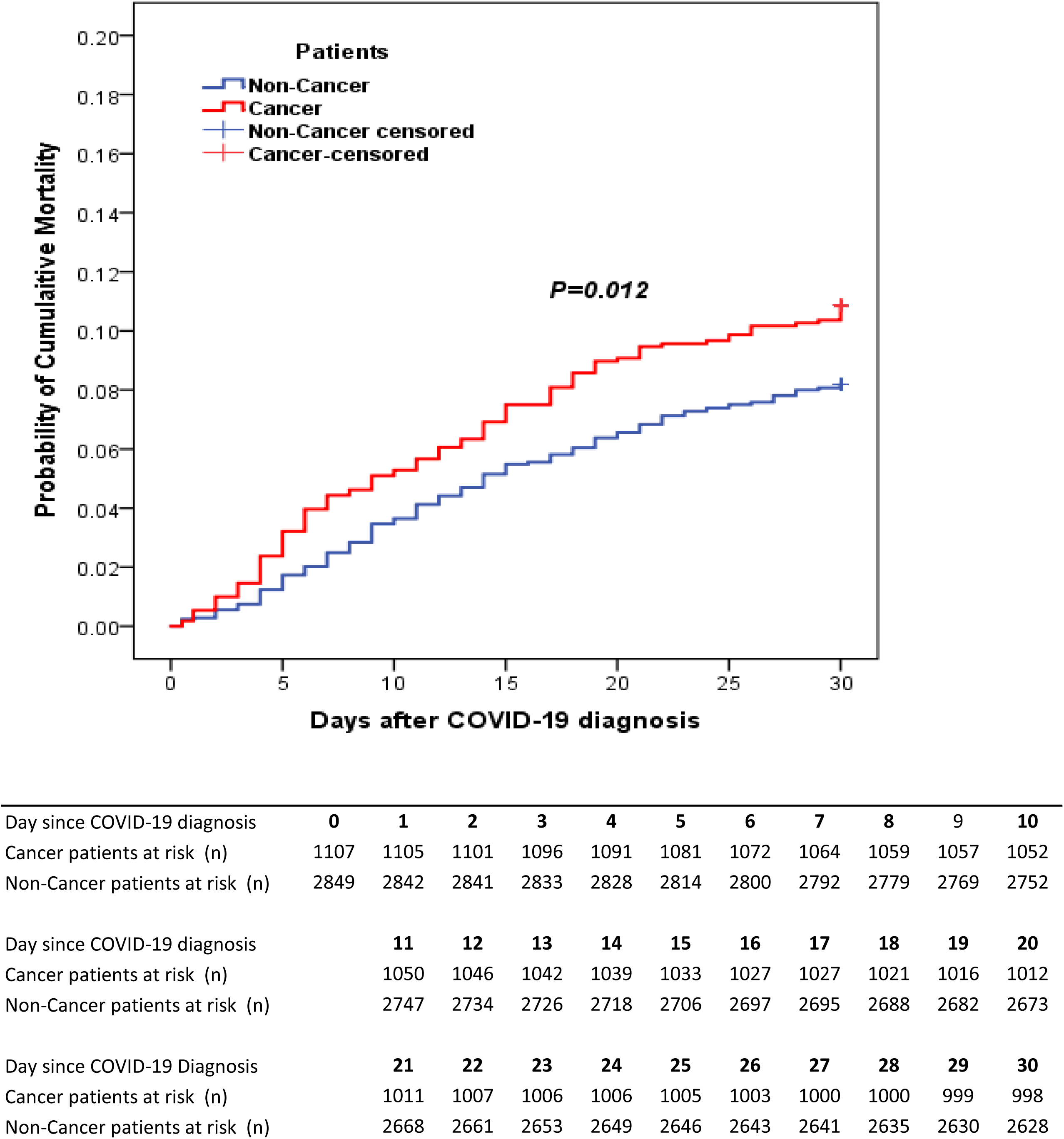
Kaplan-Meier mortality curves of COVID-19 patients with and without cancer (N=3956).

### Risk factors for death within 30 days after COVID-19 diagnosis

Of the total 3966 patients 348 died within 30 days (8.8%) and of them 189 had data available for their cause of death, which showed 93% (176/189) of the deaths were COVID-19 related. In the multivariable logistic regression analysis including all patients, cancer was an independent risk factor for 30-day mortality (OR 1.46; 95% CI 1.03 to 2.07; p=0.035) even though cancer patients were less frequently hospitalized and less frequently received corticosteroids and remdesivir (Figure 2). Older age (≥65 years) was the strongest predictor of 30-day mortality in all patients (OR 4.55; 95% CI 3.34 to 6.20; p<0.0001) and in both cancer and non-cancer patients (Figures 2 & 3).

**Figure 2.**
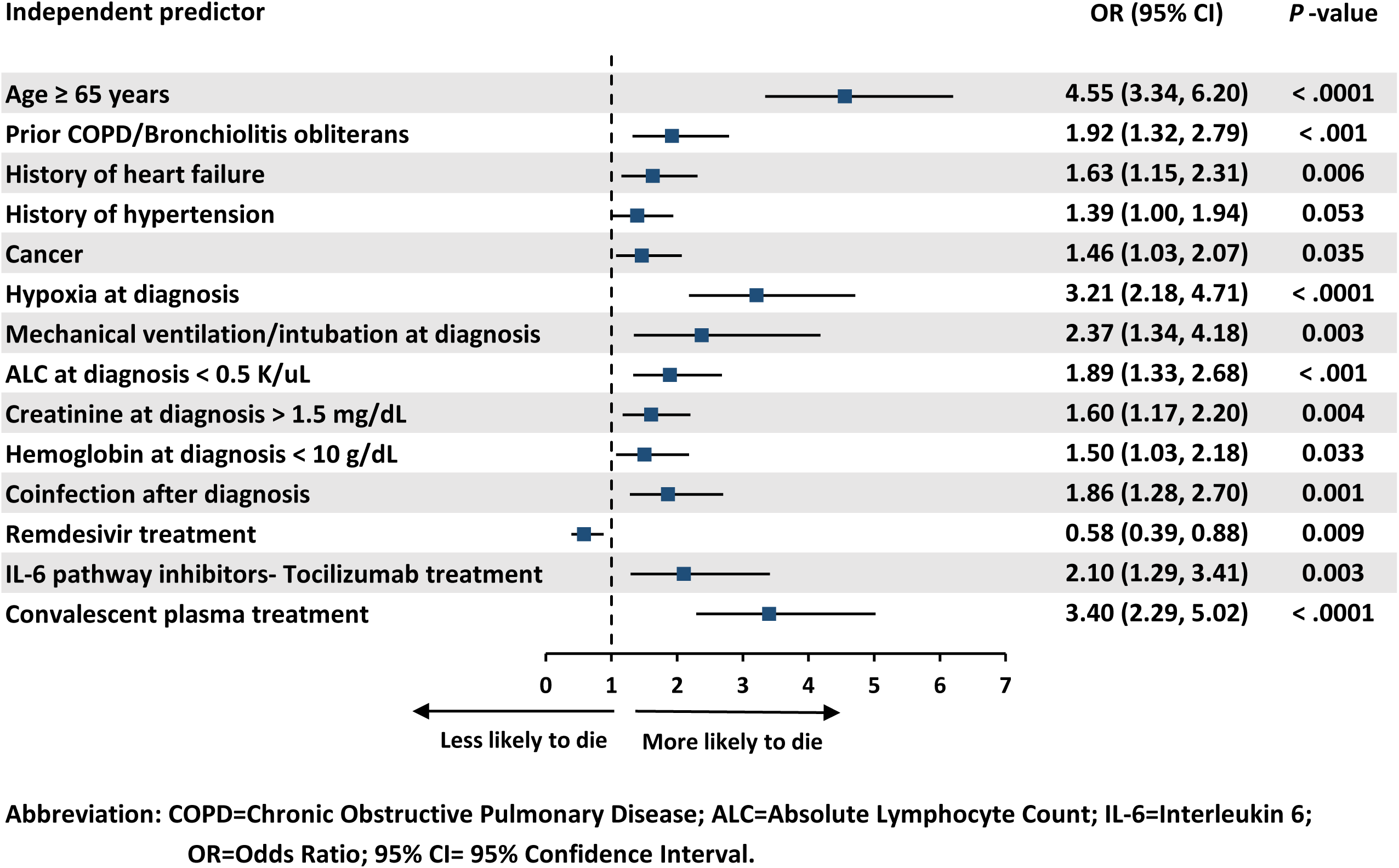
Independent predictors of 30-day mortality of COVID-19 patients by multivariable logistic regression analysis (N=2349).

**Figure 3.**
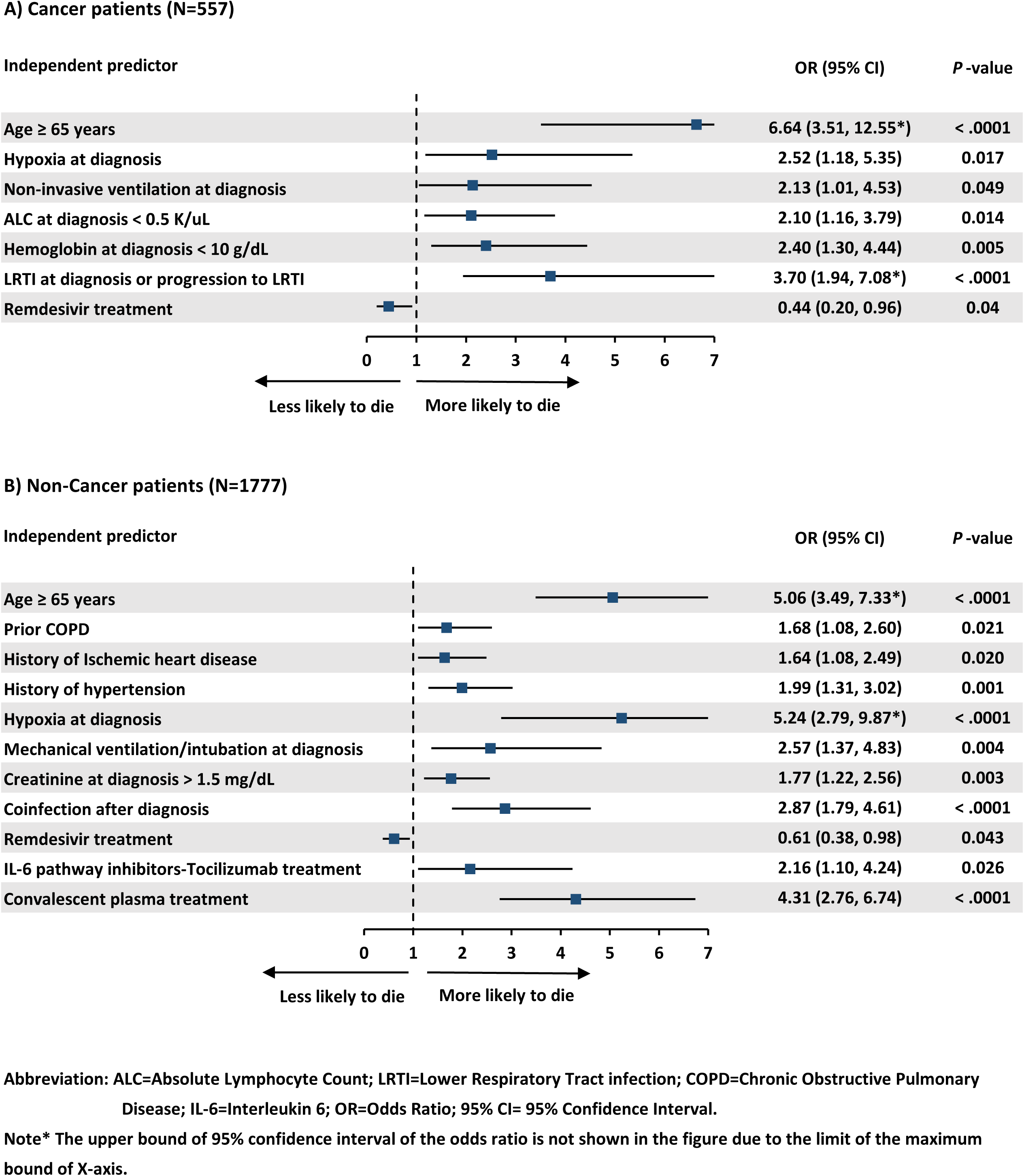
Independent predictors of 30-day mortality of COVID-19 patients with cancer (A) and without cancer (B) by multivariable logistic regression analysis.

Other independent risk factors for 30-day mortality in all patients included hypoxia at diagnosis (OR 3.21; 95% CI 2.18 to 4.71; p<0.0001), need for mechanical ventilation (OR 2.37; 95% CI 1.34 to 4.18; p=0.003), and presence of co-infection (OR 1.86; 95% CI 1.28 to 2.70; p=0.001) (Figure 2).

In cancer patients, lower respiratory tract infection manifested by the presence of pulmonary infiltrates either at diagnosis or during the course of infection was a strong independent predictor of 30-day mortality (OR 3.70; 95% CI 1.94 to 7.08; p<0.0001) (Figure 3). Among cancer patients, the 30-day mortality rate was significantly higher in patients with lung cancer than in patients with non-lung cancer solid tumors, including those with lung metastases (p<0.001; Table 2). Patients with hematologic malignancies had a significantly higher 30-day mortality than patients with non-lung cancer solid tumors (p<0.001) but tended to have a lower mortality rate than patients with lung cancer (p=0.07; Table 2). We did not find a significant difference in mortality between hematological malignancies and solid tumors by multivariate analysis (p=0.30).

**Table 2.**
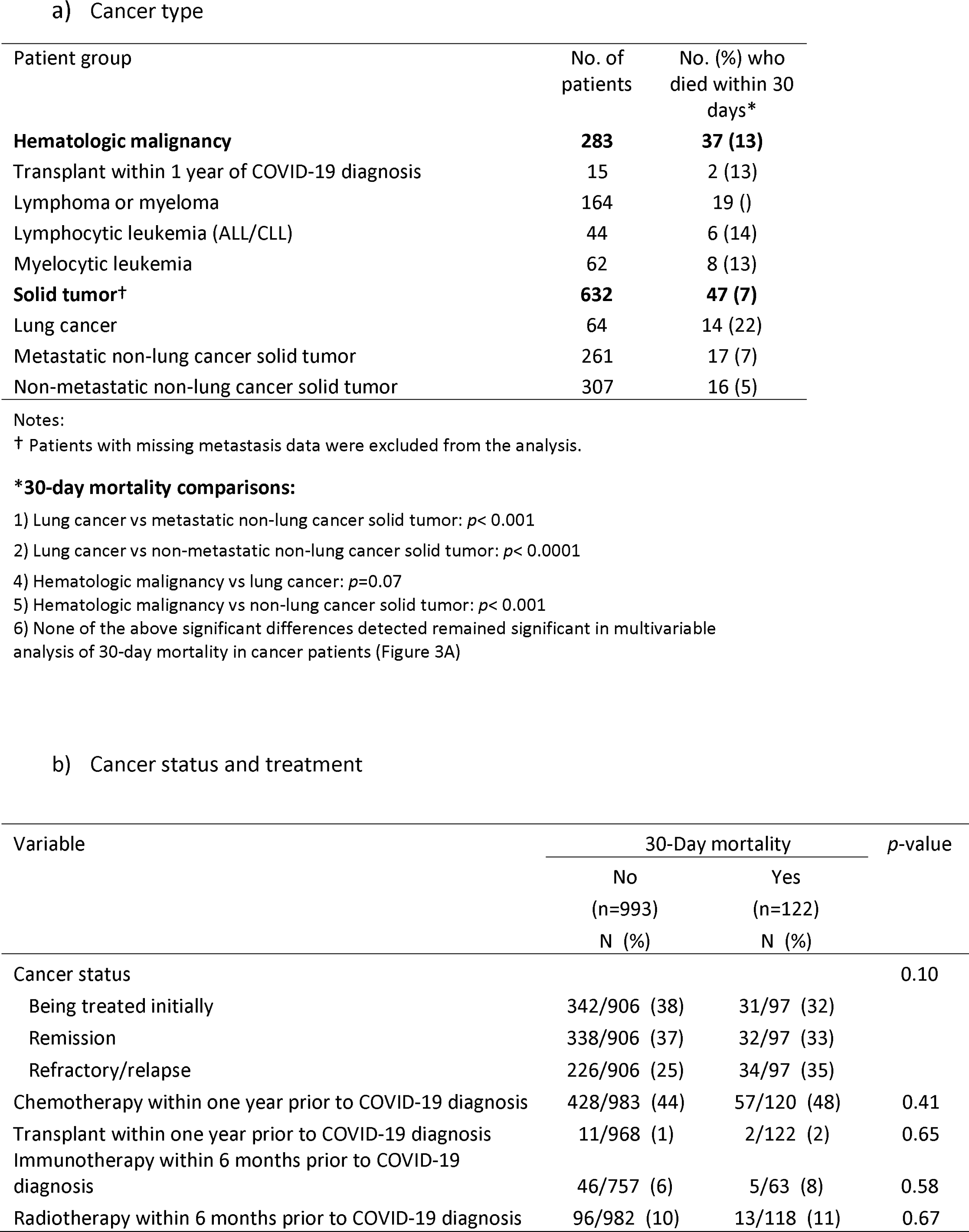
30-Day mortality among different groups of cancer patients with COVID-19

By multivariate analysis, remdesivir was the only therapeutic agent independently associated with decreased 30-day all-cause mortality in all patients (OR 0.58; 95% CI 0.39 to 0.88; p=0.009) (Figure 2), in cancer patients (OR 0.44; 95% CI 0.20 to 0.96; p=0.04) and non-cancer patients (OR 0.61; 95% CI 0.38 to 0.98; p=0.04) (Figure 3).

Among patients on low-flow oxygen at admission, the mortality rate was lower among those who received remdesivir than among those who did not (5.9% [3 of 51] vs 17.6% [68 of 387]; p=0.03). However, among patients on high-flow oxygen at admission, there was no difference in the mortality rate between patients who received remdesivir and those who did not (29.7% [11 of 37] and 34.4% [53 of 154], respectively; p=0.59).

Since 85% of patients treated with remdesivir also received corticosteroids, we evaluated the impact of their combination therapy on 30-day mortality. Among patients on low-flow oxygen at admission, the mortality rates among those who received remdesivir alone or in combination with corticosteroids (0% [0 of 8] and 7.0% [3 of 43], respectively) tended to be lower than the mortality rate of those who received corticosteroids alone (18.3% [21 of 115]; p=0.10). However, mortality rates were similar for remdesivir alone and combination therapy among various patients’ groups (Table 3).

**Table 3.** Comparing Mortality in Patients Treated with Remdesivir Alone or with Steroids for COVID-19.

| Patients groups | Outcome | Remdesivir and<br>steroids | Remdesivir alone | *p*-value |
| --- | --- | --- | --- | --- |
| All patients | 30-Day death | 62/370 (16.8%) | /65 (18.5%) | 0.74 |
| Patients with Hypoxia | 30-Day death | 58/336 (17.3%) | 10/53 (18.9%) | 0.77 |
| With low-flow oxygen | 30-Day death | 3/43 (7%) | 0/8 (0%) | >.99 |
| With high-flow oxygen | 30-Day death | 5/18 (27.8%) | 6/19 (31.6%) | 0.8 |
| On ventilation at diagnosis | 30-Day death | 26/146 (17.8%) | 7/30 (23.3%) | 0.48 |

We then evaluated the impact of timing administration of different agents on 30-day mortality. We found that patients who died within 30 days were less likely to be transfused with convalescent plasma within 1 day of COVID-19 diagnosis (which likely occurs 2-3 days after symptom onset) than patients who survived (1% vs 7%, *p*=0.04; Supplemental Table 1). Giving convalescent plasma to patients later than 3 days after diagnosis did not make any difference in mortality (Supplemental Table 1). In contrast to what was observed for convalescent plasma, the later corticosteroids were administered, the greater the benefit. There was a trend toward a greater reduction in 30-day mortality in patients who received corticosteroids later (>5 days after diagnosis), whereas no difference was observed in patients who received corticosteroids earlier.

## DISCUSSION

Unlike the previously published studies that compared patients with COVID-19 with and without cancer, our study included a large number of cancer patients from five different continents. Our findings demonstrate that cancer is an independent risk factor for increased 30-day all-cause mortality, which is consistent with a recent large study in the US that also showed that patients recently receiving cancer treatment had a worse outcome.^19^ Furthermore, this higher mortality rate in cancer patients seems to be driven by patients with lung cancer and patients with hematologic malignancies, a finding that is consistent with prior literature.^8,9^ The mortality rate in the patients with solid tumors other than lung cancer was not different from the mortality rate in the non-cancer patients. Most of the other independent risk factors that we identified for 30-day mortality after a diagnosis of COVID-19 have also been commonly reported as risk factors for mortality in previous studies of COVID-19 in cancer and non-cancer patients.^2-13^ In addition to a higher 30-day mortality rate, the cancer patients in our study had a more complicated course after discharge from the hospital; specifically, they more frequently required supplemental oxygen and readmission within 30 days after COVID-19 diagnosis.

The presentation pattern of cancer patients with COVID-19 was different to that of non-cancer patients. Cancer patients seemed to be less symptomatic. This could be related to the fact that cancer patients tended to be older, with fewer inflammatory cells (neutrophils and lymphocytes), and more often on corticosteroids. Levels of inflammatory biomarkers were also higher in patients with cancer, particularly D-dimer, ferritin, and pro-calcitonin.

On multivariate analysis, the only therapeutic agent that independently decreased 30-day all-cause mortality in both cancer patients and non-cancer patients was remdesivir. Upon further analysis, remdesivir was found to further decrease mortality in patients with pneumonia and mild hypoxia who were receiving low-flow oxygen (≤6 liters per minute) and not in patients with severe advanced pneumonia who were receiving high-flow oxygen and/or ventilatory support. In a large multicenter prospective randomized placebo-controlled trial (ACTT-1) evaluating remdesivir in 1062 patients with COVID-19 pneumonia, this agent was found to be associated with a quicker recovery than placebo.^14^ In a further subgroup analysis, remdesivir was found to significantly reduce the time to recovery and 28-day mortality in patients who were on low-flow oxygen at baseline but not in patients who were on mechanical ventilation or extracorporeal membrane oxygenation at baseline.^14^ Hence, the results of this large prospective randomized study, particular the results of the subgroup analysis, are consistent with our findings.In a meta-analysis that examined four large prospective randomized trials, remdesivir was shown to be associated with reduced 14-day mortality and reduced need for mechanical ventilation,^20^ which also supports our analysis.

In contrast, the World Health Organization (WHO)-- sponsored multinational Solidarity Trial of COVID-19 hospitalized patients who were randomly assigned to either remdesivir (2750 patients) or standard of care (2708 patients) showed no difference in overall 28-day mortality.^21^ However, in two large meta-analyses that each examined more than 13,000 COVID-19 patients from randomized and non-randomized studies, including the WHO randomized trial, remdesivir was associated with a significant improvement in the 28-day recovery rate.^22,23^ Furthermore, in a large study evaluating treatments and outcomes among cancer patients with COVID-19, remdesivir alone was significantly associated with a lower 30-day all-cause mortality rate than other treatments (including high-dose corticosteroids and tocilizumab).^4^ In addition, in a large multicenter matched controlled study involving mostly noncancer patients, Mozaffari E et al. demonstrated that remdesivir, was significantly effective in reducing mortality in patients on low-flow oxygen but not patients with advanced disease on high-flow oxygen and ventilator support.^24^ More recently a prospective randomized, double blind, placebo-controlled study (involving mainly non-cancer patients) showed that early initiation of remdesivir prevents progression to severe COVID-19.^25^

Therefore, the cumulative data in the literature do support our finding that remdesivir in all patients (cancer and non-cancer patients) improves outcome and may reduce mortality in patients with COVID-19, particularly in patients with moderate pneumonia who are receiving low-flow oxygen. Such patients are in stage II of the three-stage COVID-19 classification previously proposed by Siddiqi et al.^26^ In that classification, stage I is mild illness and high-level viral replication that all patients go through and most (>80%) recover from. It is during this stage, where the new antiviral agents (Ritonavir – Boosted Nirmatrelvir and Molnupiravir) were found to be highly effective in reducing the risks of hospitalization and death among high risk patients with COVID-19.^27,28^ Stage II is moderate pneumonia, which includes early hypoxia necessitating low-flow oxygen. Patients with stage II COVID-19 benefit from early remdesivir since viral replication continues in association with host inflammatory response in accordance with our data, the literature, and the Siddiqi et al therapeutic model. Stage III is the hyperinflammatory phase with severe pneumonia and extrapulmonary system manifestations. Patients with disease that has progressed into this hyperinflammatory late stage with severe pneumonia and hypoxia requiring high-flow oxygen and ventilatory support do not seem to benefit from remdesivir or other antiviral therapy but could benefit from anti-inflammatory agents such as corticosteroids.

The use of corticosteroids (dexamethasone 6 mg/day) was shown in a large randomized open-label study conducted in the United Kingdom to be associated with reduced 28-day mortality, particularly in patients requiring oxygen supplementation and invasive ventilation who receive corticosteroids after 7 days from the onset of symptoms.^18^ Subsequently, two meta-analyses of several prospective randomized trials demonstrated that corticosteroid use was significantly associated with a decrease in COVID-19 mortality.^29,30^ In our study, we found that if corticosteroids were started more than 5 days after the PCR diagnosis of COVID-19, there was a trend towards a reduction in 30-day COVID-19 mortality compared to starting corticosteroids earlier. Five days after diagnosis by PCR testing would possibly be equivalent to 7 days after the onset of symptoms since the average time from symptom onset to PCR diagnosis has been estimated to be 2 to 3 days.^31^ Furtheremore, by multivariate analysis and upon further subanalysis (Table 3), remdesivir was found to improve outcome independently of the effect of steroids.

With regard to convalescent plasma treatment, multivariate analyses from our study did not show any overall benefit. This finding is consistent with a meta-analysis of randomized controlled trials comparing convalescent plasma to standard of care.^17^ However, when convalescent plasma was given very early after diagnosis (within 1 day of COVID-19 diagnosis, which is estimated to be around 3 days after symptom onset), this intervention was associated with a significant reduction of COVID-19-related 30-day mortality compared to the late intervention. Several studies have shown that early transfusion (within 3 days of hospital admission) of high-titer plasma may be associated with an improved clinical outcome of COVID-19.^15,16,32,33^ The lack of benefit noted in several studies of convalescent plasma could be related to delayed administration of convalescent plasma, in the advanced hyperinflammatory stages of COVID-19, rather than administration in the early stage when viral replication is at its highest and neutralizing antibodies in convalescent plasma could be most effective, as previously noted by Siddiqi et al., early during the infection.^15-18,20-23,26^

Our study has several limitations. First, the retrospective design precluded complete assessment of disease progression in the outpatients, which limited their input into the general study. Second, some non-cancer centers contributed data only on their hospitalized patients who were likely sicker. This may have biased the data towards a higher rate of hospitalization among non-cancer patients and made the data more heterogeneous. In addition, the predominance of symptomatic patients (87%) might also limit our evaluation of the impact of cancer in the whole picture of the disease. Third, there were differences in responses to COVID-19 from one geographic area to another. Patients diagnosed with COVID-19, may have been treated differently according to specific national guidelines. For example, the 30-day all-cause mortality rate was lower at centers in the United States than at centers in Latin America or Asia. This could have been related to the accessibility or early initiation of some interventions in the United States, such as remdesivir or convalescent plasma, which made data more heterogeneous. We did not evaluate the accessibility and timing of interventions in the treating facilities. Last, this study was conducted prior to the introduction of COVID vaccines and included patients who were not vaccinated and who were infected by early variants which limits the generalizability of our results to contemporary COVID-19 patients.

In conclusion, this is the largest multicenter worldwide study comparing cancer patients to non-cancer patients with COVID-19. In this study, although the limited effect size, underlying malignancy was found to be an independent risk factor for a higher 30-day all-cause COVID-19 mortality, with lung cancer and hematologic malignancies associated with the highest risks for poor outcome. Finally, remdesivir stood out as the only therapeutic agent independently associated with decreased 30-day mortality, particularly in patients on low-flow oxygen. Corticosteroids tended to be most useful if given more than 5 days after COVID-19 diagnosis. The role of these therapeutics and their timing of administration should be verified in larger studies, especially in cancer patients, who tend to have a higher degree of immunosuppression, which may lead to prolongation of the viral phase.

## Data Availability

The study protocol, statistical analysis plan, lists of deidentified individual data, generated tables and figures will be made available upon request by qualified scientific and medical researchers for legitimate research purposes. Requests should be sent to. Data will be available on request for 6 months from the date of publication. Investigators are invited to submit study proposal requests detailing research questions and hypotheses in order to receive access to these data.

## Contributors

IR, RH, AMC were responsible for study conception, protocol design, writing of the first draft and coordinating the manuscript. RH, NM, TD, HD, BS, AB, RS, AFC, EG, AB, NM, NH, SS, TD, YEVK, SF, PAH, JB, FK, BG, TJ, EYG, HT, NRA, MY, CCRG, MP. NA. RDR, SMA, RED, TM, KI, RH, RAN, AGG, CTP, DGL, MS, FT, AEM, and AMC were responsible for data collection at their respective sites. YJ designed the statistical analysis plan, analyzed the data, developed the figures and the tables. IR, RH, HD, YJ, VS, AEM, and AMC had access to all the data and were responsible for overall project and data management. All authors had full access to the study data and had final responsibility for the decision to submit for publication. All authors were responsible for critical review of drafts and approval of the final manuscript.”

## Funding

National Cancer Institute, National Institutes of Health

**Supplemental Table 1.** Timing of administration of convalescent plasma and corticosteroids as COVID-19 treatment and 30-day mortality

| Treatment and timing of initiation in relation to COVID-19 diagnosis | Alive<br>N/n (%) | Dead<br>N/n (%) | *p*-value |
| --- | --- | --- | --- |
| **Convalescent plasma** |  |  |  |
| ≤ 1 day | 14/196 (7) | 1/88 (1) | 0.04 |
| > 1 days | 182/196 (93) | 87/88 (99) |  |
| ≤ 2 days | 49/196 (25) | 17/88 (19) | 0.3 |
| > 2 days | 147/196 (75) | 71/88 (81) |  |
| ≤ 3 days | 80/196 (41) | 37/88 (42) | 0.9 |
| > 3 days | 116/196 (59) | 51/88 (58) |  |
| **Corticosteroids** |  |  |  |
| ≤ 3 days | 594/789 (75) | 131/165 (79) | 0.3 |
| > 3 days | 195/789 (25) | 34/165 (21) |  |
| ≤ 4 days | 633/789 (80) | 139/165 (84) | 0.2 |
| > 4 days | 156/789 (20) | 26/165 (16) |  |
| ≤ 5 days | 665/789 (84) | 147/165 (89) | 0.1 |
| > 5 days | 4/789 (16) | 18/165 (11) |  |
| ≤ 6 days | 700/789 (89) | 153/165 (93) | 0.1 |
| > 6 days | 89/789 (11) | /165 (7) |  |

Note: Diagnosis of COVID-19 by PCR occurs around 2-3 days after onset of symptoms.

## Acknowledgments

We thank all the investigators, MD Anderson Cancer Network, and the Data-Driven Determinants for COVID-19 Oncology Discovery Effort (D3CODE) Team at The University of Texas MD Anderson Cancer Center for assistance in study development and data extraction.

We thank Ms. Meena Medepalli and Mr. Joel Cox from the MD Anderson Cancer Network - External Research at The University of Texas MD Anderson Cancer Center for their support in coordinating the cancer network sites for protocol participation.

We thank Mr. Joseph P. Thomas at The University of Texas MD Anderson Cancer Center for his assistance with data capture in REDCap, Kris Weaver for data quality review, Sheri Rivera for REDCap build, Regulatory team for the fast activation, Toby and her team for assistance in resolving IT issues with site EMR.

We thank Anastasia Turin and Drew Goldstein from Syntropy Technologies LLC, Burlington, MA for interfacing with the Syntropy platform: Palantir Foundry. “Foundry” is Syntropy’s fully-managed, cloud-based software-as-a-service for governing, structuring, and harmonizing real-world data to empower health systems and their collaborators to derive insights from that data.

Editorial assistance was provided by Stephanie P Deming, Research Medical Library at MD Anderson. This assistance was funded by The University of Texas MD Anderson Cancer Center.

We thank Ms. Salli Saxton and Ms Christine Cobb at The University of Texas MD Anderson Cancer Center for helping with the submission of the manuscript.

